# Estimating cost of integrating HBV, HCV, and HIV screening at Antenatal using Time-Driven Activity Based Costing (TDACB) Approach; A provider’s perspective comparing Intervention and standard of care at lower health facilities in West Nile sub region, Uganda

**DOI:** 10.64898/2026.05.20.26353753

**Authors:** John Bosco Alege, John Paul Oyore, Rose Clarke Nanyonga, Anthony Ssebagereka, Richard Ssempala, Philippa Musoke, Alloys S.S Orago

## Abstract

*Objective:* To Estimate cost of integrating HBV, HCV, and HIV screening at Antenatal using Time-Driven Activity Based Costing (TDACB) Approach; A provider’s perspective comparing Intervention and standard of care at lower health facilities in West Nile sub region, Uganda

*Methods:* Design
The Time Driven Activity-Based Costing (TDABC) approach was used to capture resource use and costs associated with delivering integrated HBV, HCV, and HIV screening among pregnant women. This study compared screening uptake among study participants in the intervention, and control group respectively. Five lower health facilities in Koboko and Maracha districts respectively in West Nile region of Uganda. A total of 1,338 study participants wo were pregnant mothers in first ANC, first trimester at the selected 10 facilities were enrolled in this study. Data were abstracted, and also collected on; Personnel/staff time; facility space utilisation; and Medical and non-medical equipment. Total cost per patient visit=Staff time costs+Space cost Equipment cost. Outcome Measure was the estimated provider-perspective costs of delivering integrated screening for HBV, HCV and HIV, using Integrated Care Model by comparing intervention and control groups. Results
Staff CCRs demonstrated considerable variability across cadres and facilities, with an overall mean of USD 0.492 per minute (Range: USD 0.167 - 1.318). Laboratory technicians exhibited the highest mean CCR at USD 0.767 per minute for personnel CCRs per patient visit. the mean lowest CPP visit was noted for HBV in the intervention arm (USD 11.43) while HIV test was the lowest in the control arm (USD 0.43). HCV test had the highest cost in the control arm (USD 0.52). The CPP visit for positive clients were generally higher than those that were negative. Equipment CCRs were minimal and highly consistent across facilities, with a mean of USD 0.00069 per minute (±0.0002). HIV/Syphilis combo was the costliest test kits at USD 3.14 per test kit followed by viral hepatitis C test kit and Hep B at USD 2.47 and USD 0.28 respectively. Facility space CCRs exhibited moderate variation across facilities, ranging from USD 0.01593 to USD 0.03474 per minute. Overall mean CCR for the space for delivering HBV, HCV or HIV testing was USD 0.0256 (0.0066). Conclusion
Overall, the integration of screening resulted in: Cost efficiencies where the same staff and space were used for multiple simultaneous tests, reduced marginal costs for HIV tests due to larger procurement volumes, and higher marginal cost additions for HBV and HCV due to pricier reagents.

## Introduction

Human immunodeficiency virus (HIV), one of the most common blood-borne viruses, has overlapping modes of transmission with the two hepatotropic viruses, hepatitis B virus (HBV) and hepatitis C virus (HCV) [1]. These two viruses (HBV & HCV) are the most common viral etiologic agents of chronic liver disease (CLD). AIDS-related illnesses have been responsible for the death of 36.3 million people since the epidemic started,[2]. In comparison, hepatitis B and C infections are accountable for 90% of viral hepatitis-related deaths (1.4 million) per annum globally [3]. All the three diseases have similar modes of transmission [4]. Several interventions have been rolled out to prevent and control new infections of the three infectious diseases (HBV, HCV and HIV) including establishing the cost of each or a combination of interventions as single diseases. Moreover, integration of disease management of similar infections and burden has been recommended as an efficient and cost-effective strategy to expand access to services [5]. Reviews, evaluation and synthesis of available literature on costing viral infectious disease services at lower health facility level have yielded various results. The numerous studies have used several costing approaches, frameworks, models and methods. For instance, studies conducted in Cambodia used Cost-effectiveness evaluation [6]. While in an evaluation conducted in the Netherlands the Markov transition state model was developed and used [7]. A systematic review conducted in Sub-Saharan Africa documented the use of “Bottom-up and step-down” costing methods to estimate cost [8-9]. In Uganda the Time-Driven Activity-Based Costing (TDABC) approach was used to conduct a study on estimating costs and resource distribution of direct services for HIV alone in selected regions of the country [10]. The study observed a mean cost of HCT services at US$8.18 per visit. Nevertheless, little is known about the cost of delivering integrated HBV, HCV and HIV care using the (TDABC) model; and whether integration increases the uptake of HBV, HCV and HIV screening at ANC. This study in Westline makes the first attempt to estimate the cost of integrated screening of HBV, HCV, and HIV at one point of care (ANC) by comparing the intervention group and standard care package. Thus, the need to carry out this study to make a case for investment/funding, prioritization and priority setting by MoH, health partners and diagnostic companies while responding to the three infectious diseases (HBV, HCV, and HIV).

## Methods

### Study setting

This was a facility-based costing study which used a Time-Driven Activity-Based Costing (TDABC) model. This model measures costs at the patient directly observing resources allocated throughout the patient’s care visit, and is applied at the facility level. Developed by Robert Kaplan and Michael Porter at Harvard Business School Robert [11-12]. Unlike previous versions of activity-based costing, TDABC employs a time estimate for the utilization of each resource to allocate costs to services in which process maps were also developed [13]. Refer to (Annex: 1 Attached as additional file) for details of the sample process maps.

### Sample, eligibility and consent

This study was conducted at 10 lower health facilities delivering integrated screening for Hepatitis B Virus (HBV), HIV, and Hepatitis C Virus (HCV) within antenatal care (ANC) in Koboko and Maracha districts, West Nile Region respectively. The facilities were grouped in to two, namely the Intervention arm, and the Control. Facilities that were grouped into the Intervention arm include; Ayipe HCIII, Dranya HCIII, Dricile HCIII, Gborokolongo HCIII, and Lobule HCIII. While those in the Control arm include; Eliofe HCIII, Kamaka HCIII, Kijomoro HCIII, Nyadri HCIII, and Oleba HCIII). TDBAC was used since it incorporates time as a critical dimension in resource allocation and costing, allowing for a more dynamic assessment of resource utilization during service delivery over varying periods. This is particularly suited to healthcare settings where activities fluctuate based on patient flow and service demands. This study aimed to estimate the cost of integrated screening for HBV, HCV, and HIV at ANC using rapid diagnostic test kits. Study participants were pregnant mothers screened for HBV, HCV and HIV in first ANC visit, first trimester at the selected health facilities in Koboko and Maracha districts. While the healthcare workers and administrators were key informants.

### Study Procedure

This study was part of a larger *Quasi* experimental study that evaluated the effectiveness of integrated viral hepatitis B, C and HIV Care Model to Optimize screening among pregnant at health facilities in West Nile region, Uganda. The ANC screening pathway was mapped from patient arrival to exit, with emphasis on activities related to HBV, HCV, and HIV screening. All the core steps for conducting the screening for the three viral infections were assessed (Reception, Registration, Health education, pre-test counselling, Sample collection, Testing, Interpretation of results and communication of results, and post-test counselling and linkage). For each service delivery step above, the following were identified: The cadre(s) involved (midwifes, counsellor, and community health worker), the physical space used (ANC room, laboratory, waiting area), the equipment required and used were (lancets, sharps, and testing devices), consumables (HBV, HCV, and HIV rapid diagnostic kits, alcohol swaps, and other supplies), and Time spent (in minutes) by each cadre delivering a service at each step.

### Description of costing screening pathways

Estimation of Capacity Cost Rates (CCRs); in order to compute the unit cost per minute of using each resource cost category (done per cadre, equipment, and facility space), the CCRs were calculated as follows:

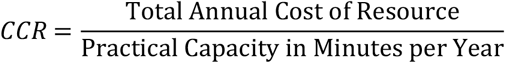

Calculation of Client-Level Costs per patient visit; For each patient visit type (HBV, HCV, HIV testing) and for each facility, we allocated costs to each patient shadowed. We multiplied the relevant CCRs by the time (in minutes) that each resource was used per patient. For each facility and test type (HBV, HCV, HIV), total cost per patient visit was calculated as follows:

Total cost per patient visit=Staff time

costs+Space cost Equipment cost

### Resources and Cost Inputs

TDABC approach was used to capture the resource use and costs associated with delivering integrated HBV, HCV, and HIV screening during ANC visits by pregnant women in their first trimester. Data were abstracted from facility reports, facility records including human resource records, facility-level and district-level key informant interviews. Data were collected on the following cost parameters based on these resource categories: Personnel/staff time: Staff salaries, allowances, working schedules, public holidays, work-related leave, and clinical time spent attending to each client; facility space utilisation: Clinic room sizes, annual maintenance costs, and hours of availability (facility operational time); Medical and non-medical equipment: Purchase price, useful life, annualized replacement value, and annual operating costs; And, test kits and reagents (for HBV, HCV, and HIV tests): unit prices, quantities used per test.

Practical capacity was computed and preferred over theoretical capacity to better accurately capture service availability of the resource. The practical capacity of each resource was calculated as the total number of minutes per year during which the resource was realistically available for patient care. This accounted for: Clinic operating hours, Equipment uptime versus downtime, Facility opening schedules, and Shared spaces such as; rooms used for multiple purposes). For each resource (staff, space, equipment), we estimated the *practical capacity* in minutes per year, accounting for: Staff capacity: Based on official working hours, number of working days per year, and adjustments for leave, meetings, and non-productive time (e.g. administrative duties)

Space capacity; For each ANC or testing room, we estimated-Hours the room was available and usable per day, Days per year the service was available, and Computed room-minutes per year

Equipment capacity; For the diagnostic and clinical equipment used, we considered-Expected useful life (years), Annual operating hours, and Practical capacity in minutes per year

The rationale as well the advantage of choosing the TDABC approach compared to more traditional costing methods in healthcare settings such as the “bottom-up” approach and others is that process maps, and or client flow charts are developed. Such maps, and or charts provide insight into how the screening for HBV, HCV and HIV was delivered. The approach also allows for easy comparisons between clients and facilities, takes a shorter time and is cost [14]. This study aimed to estimate the cost of integrated screening for HBV, HCV, and HIV at ANC using rapid diagnostic test kits.

### Costing perspective and approach

The costing model involved only health facility inputs. Resources associated with both the intervention and control group pathways were measured through observation of standard operating procedures as health workers and supporting staff performed their duties in respective sections within the pathway. Staff salary, for example, was allocated based on the time spent on the reference case (described in the assumptions) as a proportion of monthly worktime. The useful life span of medical equipment used was considered according to the manufacturers’ instructions where possible or an estimated period from expert opinion. Building space occupied was given an assumed expected lifetime of 30 years. All costs were estimated as of mid-year prices of 2025 and converted to US dollars using published Bank of Uganda exchange rate of US$1US D=UGX3700. The time horizon for cost analysis was 1 year and thus discounting for future costs was not done. We also varied the costs of consumables because their values were majorly obtained from expert opinion and fluctuating market prices.

We used the Consolidated Health Economic Evaluation Reporting Standards (CHEERS) reporting guidelines [30] to verify and ensure standard reporting of this research work. The study assumed that all the overhead costs of infrastructure, RDTs, laboratory tests, sample collection requirements and biosafety requirements were the same or similar for the intervention and control group. The useful life of the medical equipment was assumed to vary between 2 and 5 years. The useful life of the furniture in the clinics was assumed to vary between 5 and 10 years depending on the type and 100% of that use was allocated to patient use. This assumption may have affected the overall accuracy of the cost estimates made in this study.

### Data collection and analysis

A Process mapping guide, Inventory of resources (personnel, the place-health facility, building surface area, and Patient time sheet, and lastly an Observation checklist were the data collection tools used in this study.

Cost estimates were obtained from clinic inputs and procurement invoices. Additional information was obtained from budgetary documentation reviews, procurement guides, and publicly available product information. Expert opinion was sought from suppliers, local distributors, and health workers. Previous costing studies were reviewed to validate some of the estimates [9]. More cost data were obtained from the health facility administration and accounts department, health facility records such as delivery notes, budgets, and invoices among others. The approach to the collection and analysis of the data is described in the following sections:

Step One: Defined the care pathways and activities; The ANC screening pathway was mapped from patient arrival to exit, with emphasis on activities related to HBV, HCV, and HIV screening. All the core steps for conducting the screening for the three ailments were assessed (Reception, Registration, Health education, pre-test counselling, Sample collection, Testing, Interpretation of results and communication of results, and post-test counselling and linkage). For each service delivery step above, the following were identified:

The cadre(s) involved (midwifes, counsellor, and community health worker); The physical space used (ANC room, laboratory, waiting area);The equipment required and used were (lancets, sharps, and testing devices); Consumables (HBV, HCV, and HIV rapid diagnostic kits, alcohol swaps, and other supplies); Time spent (in minutes) by each cadre delivering a service at each step.

Annual capacity (Practical Capacity) for each resource

Practical capacity was computed and preferred over theoretical capacity to better accurately capture service availability of the resource. The practical capacity of each resource was calculated as the total number of minutes per year during which the resource was realistically available for patient care. This accounted for: Clinic operating hours, Equipment uptime versus downtime, Facility opening schedules, and Shared spaces such as; rooms used for multiple purposes). For each resource (staff, space, equipment), we estimated the *practical capacity* in minutes per year, accounting for:

Staff capacity: Based on official working hours, number of working days per year, and adjustments for leave, meetings, and non-productive time (e.g. administrative duties)

Space capacity: For each ANC or testing room, we estimated:

- Hours the room was available and usable per day
- Days per year the service was available
- Computed room-minutes per year

Equipment capacity: For the diagnostic and clinical equipment used, we considered:

- Expected useful life (years)
- Annual operating hours
- Practical capacity in minutes per year

Step Two: Estimation of Capacity Cost Rates (CCRs)

In order to compute the unit cost per minute of using each resource cost category (done per cadre, equipment, and facility space), the CCRs were calculated as follows:

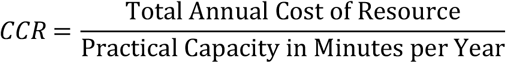

Calculation of Client-Level Costs per patient visit

For each patient visit type (HBV, HCV, HIV testing) and for each facility, we allocated costs to each patient shadowed. We multiplied the relevant CCRs by the time (in minutes) that each resource was used per patient. For example:

i. Staff time costs = (CCR for personnel × minutes spent per patient visit for Medical Diagnostic test)
ii. Space costs = (CCR for space × duration client occupied the room during the service)
iii. Equipment costs = (CCR for equipment × equipment-minutes used)

For each facility and test type (HBV, HCV, HIV), total cost per patient visit was calculated as follows: Total cost per patient visit=Staff time costs+Space cost Equipment cost

### Statistical analysis

We gathered descriptive statistics on service characteristics, including frequency, quantity and distribution of costs and resources for HIV, HBV and HCV service provisions. All costs were reported in USD with data collection spanning 2024-2025.

All component costs were aggregated to determine total cost per client for each disease screening category (HBV, HCV, HIV). Analysis outputs were summarized into descriptive statistics (means, SD, Range), and graphs were used to summarise CCRs and costs per-patient costs. We also made comparisons of the costs between the facilities in the Intervention compared to the control arm, by using ttest and chi square tests.

### Ethics

The research protocol for this study was reviewed and approved by the CIU-REC and Uganda Council for Science and Technology. Free and voluntary consent was sought from the study participants before embarking on the study. The study was conducted in accordance with the Declaration of Helsinki since human subjects were the participants. The research protocol for this study was reviewed and approved by the CIU-REC and Uganda Council for Science and Technology. Free and voluntary consent was sought from the study participants before embarking on the study.

## Results

### Respondent characteristics

A total of 1338 pregnant mothers in first trimester participated in this study, out of whom, 705 were allocated to the intervention arm while 633 participants were in the Control arm. Study participants were enrolled across ten (10) Health Centre III (HC III) facilities in the West Nile Region. A total of 705 (52.7%) were allocated to an integrated screening, which was also the intervention arm, while 633 (47.3%) received standard care package, and was the (Control arm). All the ten (10) facilities were public HC IIIs, providing a homogeneous facility-level context for analysing cost drivers related to integrated screening for HBV, HCV and HIV.

**Cost of integrated screening of HBV, HCV, and HIV screening services at ANC using Time Driven Activity Based Costing (TDABC) were estimated as shown in Table 1**.

**Table 1:**
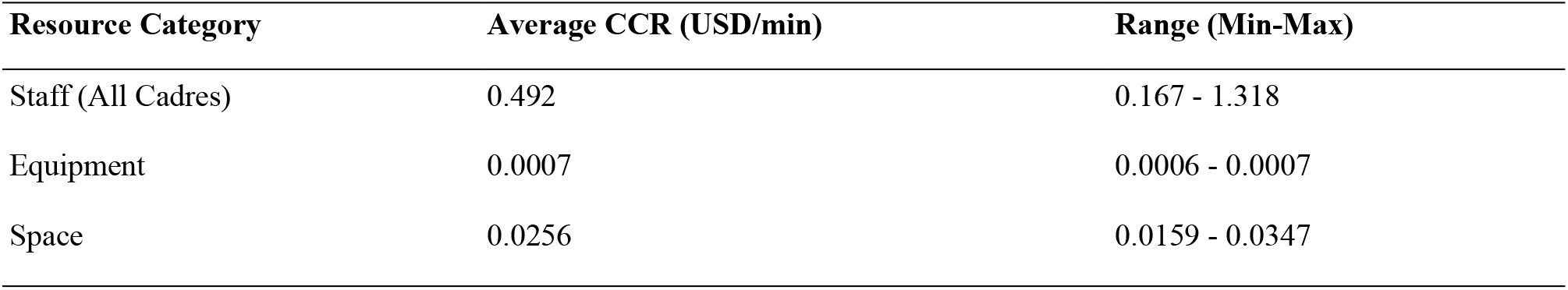
Summary of average CCRs by resource category across facilities.

| Resource Category | Average CCR (USD/min) | Range (Min-Max) |
| --- | --- | --- |
| Staff (All Cadres) | 0.492 | 0.167 - 1.318 |
| Equipment | 0.0007 | 0.0006 - 0.0007 |
| Space | 0.0256 | 0.0159 - 0.0347 |

### Summary of Average CCRs by Category across facilities

**Table 2(a):** Mean staff Capacity cost rates (in USD/min) by intervention type.

| Intervention type | Mean CCR | SD |
| --- | --- | --- |
| Control | 0.45 | 0.16 |
| Intervention | 0.53 | 0.33 |
| **Overall** | **0.49** | **0.26** |

### Staff capacity cost rates and cost per patient visit

Staff CCRs demonstrated considerable variability across cadres and facilities, with an overall mean of USD 0.492 per minute (Range: USD 0.167 - 1.318) as shown in (Table *1*) above. This variation reflects differences in annual salary scales, staffing patterns and the practical capacity (available patient-contact time) of staff members across the facilities.

The analysis revealed seven key staff categories involved in integrated viral hepatitis and HIV screening at ANC: midwives, nurses, laboratory technicians, counsellors, and community health workers (CHWs). The staff CCRs quantify the estimates for the cost per minute to deliver a given specified service.

As detailed in Table 2(b) below, laboratory technicians had the highest mean CCR (USD 0.84 ± 0.42), followed by counsellors (USD 0.58 ± 0.05). Midwives, who form the backbone of ANC service delivery at HC IIIs, had lower but more consistent CCRs (USD 0.39 ± 0.02 and USD 0.47 ± 0.00, respectively). There was a higher CCR for the intervention arm (USD 0.53 ± 0.33) compared to the control arm (USD 0.45 ± 0.16), though not statistically significant (p=0.188). Nurse CCRs *exhibited uniformity across all ten facilities at USD 0*.*468 per minute, with no variation*.

**Table 2(b):** Mean staff Capacity cost rates (in USD/min) by cadre and intervention type.

| Cadre<sup>1</sup> | Intervention | Control | Overall |
| --- | --- | --- | --- |
|  | CCR Mean (SD) | CCR Mean (SD) | CCR Mean (SD) |
| CHW | 0.21 (0.02) | 0.22 (0.00) | 0.21 (0.02) |
| Counsellor | 0.56 (0.07) | 0.59 (0.00) | 0.58 (0.05) |
| Lab Tech | 1.14 (0.39) | 0.53 (0.00) | 0.84 (0.42) |
| Midwife | 0.39 (0.03) | 0.38 (0.00) | 0.39 (0.02) |
| Nurse | 0.47 (0.00) | 0.47 (0.00) | 0.47 (0.00) |
| **Overall** | **0.53 (0.33)** | **0.45 (0.00)** | **0.49 (0.26)<sup>2</sup>** |

<sup>1</sup> In the analysis, we excluded clinical officer and receptionist, because of minimal involvement to the process of screening and testing for HBV, HIV and HCV clients.
<sup>2</sup> $t = -1.3285$ ( $p=0.1884$ )

These variations in CCR could be due to: (1) differences in staff salaries across cadres; (2) differences in staffing norms (e.g., more staff sharing workload reduces CCR); (3) facility operating hours. Staff with generally higher CCRs indicate that they either have higher renumeration or lower available patient-contact times.

### Personnel cost capacity rates per patient visit

Laboratory technicians exhibited the highest mean CCR at USD 0.767 per minute. Higher CCRs reflect higher annual staff salaries, lower practical capacity, or both. In addition, Laboratory technician CCRs demonstrated the widest variation among all personnel categories, ranging from USD 0.446 to USD 1.318 per minute, with a mean of USD 0.767 per minute. This represents a 2.95-fold difference between minimum and maximum values—by far the largest variation observed across all staff categories. Laboratory technicians emerged as the most costly and variable personnel category, playing a critical role in specimen processing, diagnostic testing, quality control, and results reporting. For integrated HIV/hepatitis screening, laboratory technicians are responsible for specimens, performing rapid diagnostic tests, operating and maintaining laboratory equipment, and participating in quality assurance activities.

For each facility, midwives (who conduct most ANC, assessment of pregnant women, counselling, and screening tasks) contribute the largest share of staff cost per patient for HBV, HCV, and HIV screening. Midwife CCRs exhibited moderate variation, with values ranging from USD 0.379 to USD 0.448 per minute. Ayipe HCIII again showed the highest midwife CCR at USD 0.448 per minute. The remaining nine facilities uniformly maintained a midwife CCR of USD 0.379 per minute. The elevated CCR at Ayipe may indicate fewer midwives relative to patient load (reducing practical capacity), or differences in the scope of midwife responsibilities at this facility. Counsellors contribute a smaller but non-trivial component. Counsellor CCRs ranged from USD 0.435 to USD 0.593 per minute, with a mean of USD 0.578 per minute. Ayipe HCIII had the lowest counsellor CCR at USD 0.435 per minute (25% below the mean), while all other facilities maintained a uniform CCR of USD 0.593 per minute. Counsellors play a critical role in both pre-test and post-test counselling for viral hepatitis and HIV screening, addressing testing procedures, risk reduction strategies, psychosocial support, and linkage to care for positive cases. The consistency of counsellor CCRs across nine facilities (USD 0.593/min) indicates standardized compensation structures and workload patterns for this cadre.

The lower CCR at Ayipe may suggest either lower counsellor salaries at this facility, higher patient volumes enabling more efficient utilization of counselling capacity, or organizational differences in how counselling time is structured, for instance; group counselling sessions versus individual. For a typical counselling session lasting 20 minutes, costs would be USD 8.70 at Ayipe compared to USD 11.86 at other facilities—a USD 3.16 difference per patient.

CHWs contribute more to cost when community mobilization or outreach for ANC and testing is intensive. Community health worker (CHW) CCRs ranged from USD 0.167 to USD 0.217 per minute, with a mean of USD 0.212 per minute. CHWs contribute to community mobilization, health education, patient follow-up, and linkage between communities and health facilities. Ayipe HCIII had the lowest CHW CCR at USD 0.167 per minute, while all other facilities maintained a CCR of USD 0.217 per minute.

Generally, the mean lowest cost per patient visit was noted for HBV in the intervention arm (USD 11.43) while HIV test was the lowest in the control arm (USD 0.43). HCV test had the highest cost in the control arm (USD 0.52). Overall, the cost per patient visit for the clients that were positive were generally higher than those that were negative. The cost per patient visit were also higher for those that were attended to by lab technicians, counsellors, and midwives. The cost per patient visit was the lowest for those that were attended to by CHWs.

### Equipment capacity cost rates

Equipment CCRs were minimal and highly consistent across facilities, with a mean of USD 0.00069 per minute (±0.0002). The low values indicate that while essential, the shared basic equipment such as; lancet devices, and others used across all three screening tests constituted a negligible cost driver on a per-minute basis. This is because HBV, HCV and HIV RDTs were used for diagnosis in this, Cost estimation for optimizing of screening Up-take study.

Equipment CCRs demonstrated minimal variation across facilities, ranging from USD 0.00064 to USD 0.00073 per minute. Generally, facilities with older or shared equipment tend to display lower CCRs, while facilities with newly purchased equipment show higher CCRs.

Overall mean CCR for the equipment for delivering HBV, HCV or HIV testing was (USD 0.00069 (0.0002.

### Cost of the test kits (HBV, HCV, HIV)

HIV/Syphilis combo was the costliest test kits at USD 3.14 per test kit followed by viral hepatitis C test kit and Hep B at USD 2.47 and USD 0.28 respectively. Test kit costs dominate overall service costs, representing 90-98% of total per-visit costs depending on test type and facility

Generally, the facilities in the intervention arm had a higher CCR for space compared to those in the control arm.

### Space (Facility) capacity cost rates

The CCR for facility space averaged USD 0.0256 per minute (±0.0066). Facility space CCRs exhibited moderate variation across facilities, ranging from USD 0.01593 to USD 0.03474 per minute. Variations were primarily driven by differences in room utilization rates and building maintenance costs. Larger ANC rooms demonstrated economies of scale, resulting in lower space CCRs. Space costs varied considerably across facilities. Lobule HC III demonstrated the highest space CCR at USD 0.03474 per minute, more than twice that of Kijomoro HCIII (USD 0.01593/min), the lowest-cost facility.

Since the sample had only HC IIIs, it was noted that there is moderate variation in space CCRs across facilities. However, the existing variations in the CCRs for space could be attributed to differences in space utilisation (how many hours per day the rooms are actively used) and building/room size. Larger facilities, for instance; HCIII with larger ANC rooms, had lower CCRs, indicating economies of scale, as compared to those with limited space or high maintenance costs which had higher CCRs.

Overall mean CCR for the space for delivering HBV, HCV or HIV testing was USD 0.0256 (0.0066). Generally, the integration of screening resulted in:

Cost efficiencies where the same staff and space were used for multiple simultaneous tests; Reduced marginal costs for HIV tests due to larger procurement volumes,

Higher marginal cost additions for HBV and HCV due to pricier reagents

## Discussion

Findings from this study suggest that Staff Capacity Cost Rates (CCRs) demonstrated considerable variability across cadres and facilities, with an overall mean of USD 0.492 per minute. The seven key staff categories involved in integrating HBV, HCV and HIV screening at ANC were; midwives, nurses, laboratory technicians, counsellors, and community health workers (CHWs) A similar study conducted in Cambodia using TDABC to estimate the cost of HIV screening services also noted staff CCRs variability [15]. Similarly, an evaluation conducted in Mozambique using the TDABC model observed that on average, the lowest CCR by cadre was among peer educators at US$0.01 per minute. The majority of HIV services providers were health technicians and nurses with an average CCR of US$0.07 and US$0.06 per minute [16]. Both studies in Combodia and Mozambique observed that the variations were as a result of the level of effort of staff per patient served, staff salaries and benefits. It was also noted that providing HIV services using an integrated approach such as at ANC, lowered the staff CCR due to task shifting among midwives, nurses and laboratory technicians, especially at health facilities in the Control group. The similarity in variations between this study and that in Cambodia and Mozambique could be linked to differences in annual salary scales, staffing patterns and the practical capacity (available patient-contact time) of staff members across the facilities.

This study observed that laboratory technicians (LTs) exhibited the highest mean CCR at USD 0.767 per minute. Higher CCRs reflect higher annual staff salaries, lower practical capacity, or both. Similarly, it was also noted that LTs CCRs demonstrated the widest variation among all personnel categories with a mean of USD 0.767 per minute. This represents a 2.95-fold difference between minimum and maximum values, and by far the largest variation observed across all staff categories. Cost data can also be used to establish variability on time spent on the client by the health workers. Findings from this study are in line with those from another study that used the TDABC approach conducted by on costing HIV services in Uganda [17]. They found that clients who received care at a HC IV level had more time (12 more minutes) compared to those at HCIII level. Normally HC IVs will have more Laboratory scientists, and are highly qualified with higher pay compared to HC IIIs which was the setting in West Nile where this study was conducted. Thus, costs and resources for HIV care vary widely in the study sites, and throughout Uganda. Nevertheless, such variation requires careful reflection, this is because some sources of variation may be indicative of vertical and horizontal equity within the health system, while others may arise from inequities such as not meeting the staffing norms as expected and provided for by the MoH-Uganda guidelines [17]. Nevertheless, high-quality and timely cost data are critical to inform decisions regarding health services provision [18].

This study also established that for each facility, midwives contributed the largest share of staff cost per patient for HBV, HCV, and HIV screening. It was also observed that midwife CCRs exhibited moderate variation, with values ranging from USD 0.379 to USD 0.448 per minute. Ayipe HCIII a facility in the Intervention group showed the highest midwife CCR at USD 0.448 per minute. While the remaining nine facilities uniformly maintained a midwife CCR of USD 0.379 per minute [19]. In a study conducted in Tanzania personnel especially midwives and nurses who worked closely with the mother-baby pair were the key cost drivers of testing services. The common practice at lower health facilities in both Uganda and Tanzania is that midwives interact with the pregnant mothers more often compared to the other cadres right from their entry in to the health facilities till exit. Contrary to findings from this study, data from a study conducted in East and Southern Africa, documented variations with the Midwife and nurse CCRs [20]. They noted that the average annual salary for a nurse was approximately US$5,250, slightly lower than documented in Rwanda (US$6,259) and considerably lower than that documented in Zambia (US$15,736) and South Africa ($33,976US). Important to note is that not all testing personnel (midwives, nurses and laboratory technicians) spent 100% of their time on HIV services [21]. It was further argued that sharing of personnel across different services is an important element of how the integrated model has potential to increase efficiency without necessarily compromising quality of services.

Importantly, the moderate midwife CCRs exhibited in this study, particularly in Ayipe HC III, may suggest either lower counsellor salaries at this facility, higher patient volumes enabling more efficient utilization of counselling capacity, or organizational differences in how counselling time is structured, for instance; group counselling sessions versus individual. For a typical counselling session lasting 20 minutes, costs would be USD 8.70 at Ayipe compared to USD 11.86 at the other health facilities within the study sites resulting to a USD 3.16 difference per patient. The above synthesis, and evaluation of the key findings from this study and the available literature is reasonably consistent with the roles of midwives. They are at the centre of conducting most ANC, assessment of pregnant women, counselling, and screening for HBV, HCV and HIV, using the respective RDTs. Lastly, in terms of comparability between the Intervention and Control group, this study observed a higher CCR for the intervention group compared to the control group, though not statistically significant. Nurse CCRs exhibited uniformity across all ten facilities at USD 0.468 per minute, with no variation.

In this study the mean lowest cost per patient visit was for HBV in the Intervention Group (USD 11.43) while HIV test was the lowest in the control arm (USD 0.43). HCV test had the highest cost in the control group (USD 0.52). Overall, it was noted that the cost per patient visit for the clients that were positive were generally higher than those that were negative. Meanwhile the cost per patient visit were also higher for those who were attended to by laboratory technicians, counsellors, and midwives. The cost per patient visit was the lowest for patients who were attended to by CHWs. The majority of the available literature reviewed observed higher cost per patient per visit for HIV, HBV and HCV [15]. A study conducted in Cambodia documented high cost per patient per visit on HIV screening services (USD 8.92) among pregnant mothers, while the same cost almost doubled at higher health facilities. Similarly, a study conducted in Burkina Faso on integrating hepatitis B screening and ART into routine ANC found the HBcrAg-RDT strategy to be the least expensive with a total economic cost of US$3959689, compared to HBV DNA (US$6128875), HBeAg (US$4135233), and treat-all (US$4141206) [22]. Consumables (cost of HIV, HBV and HCV RDTs), and personnel salaries drove up the cost of screening [23]. In another study conducted in Southern Africa on Costs of facility-based HIV testing in Malawi, Zambia, and Zimbabwe, a relatively lower cost of delivering HTS services at US$3 per individual tested was noted. Nevertheless, this cost is still higher than the cost observed in this study. In the case of the study from Southern Africa, the differences in the number of visits per staff were the primary determinant of differences in the cost per visit. Similar findings were noted in another study conducted in Ethiopia, where Average Cost per Patient Visit by Test was (USD 5.06), which again is higher than what was found in this study [24]. Lastly, a study conducted in Uganda using TDABC, observed a mean cost of HIV testing services at US$8.18 per visit [10]. The findings is higher than the mean cost per patient visit obtained in our study.

Generally, the variation in the Average Cost per Patient Visit by Test between findings observed in this study and those from existing literature as discussed above are mainly due to; the different costing models, and study designs used. Other differences can be derived from variation in the number of patients enrolled in the study, thus, the sample size, different purchasing power of the local currencies that directly affects procurement of testing kits, and government subsidies in some instances, for diagnostics (RDTs) and other medical supplies. Important to note is that the level of health facilities did not influence the Average Cost per Patient visit by test. Facility outcome from all the literature considered in this synthesis and evaluation were findings from studies conducted in lower health facilities only. It is also important to note that the variability in all these cost components across different settings and over time, makes comparison of studies difficult. Nevertheless, such variation requires careful reflection, this is because some sources of variation may be indicative of vertical and horizontal equity within the health system, while others may arise from inequities [10].

This study found that Equipment CCRs were minimal and highly consistent across facilities, with a mean of USD 0.00069 per minute (±0.0002). The low values indicate that while essential, the shared basic equipment such as; cupboards, couches, lancet devices, and others used across all three screening tests constituted a negligible cost driver on a per-minute basis. This is because only HBV, HCV and HIV RDTs were used for diagnosis in this Cost estimation for optimizing of screening Up-take study. Generally, facilities with older or shared equipment tended to display lower CCRs, while facilities with newly purchased equipment showed higher CCRs. Findings from this study particularly on the Equipment CCRs are consistent with existing literature on similar studies. For instance, an evaluation conducted by the MoH in Mozambique established that equipment costs were a very small proportion of the total costs, only estimated at (1 percent), and so, had minimal impact on the overall unit costs [16]. On the contrary, different findings were documented in a qualitative study conducted on “implementation of integrated screening for HIV, syphilis, and hepatitis B among pregnant women in Nepal [25]. The study noted availability of HIV kits, but no hepatitis B and C testing kits, and sometimes facilities were faced with shortages. Similarly, it was reported that laboratory services including use of simple diagnostics such as RDTs were often not available, or costly to be procured for the health posts (similar to the lower health facilities such as HCIIIs and HC IIs in Uganda). Findings from another study conducted in Ethiopia corroborate with those from the study in Nepal. Equipment and infrastructure costs accounted for (74 percent) of the resources used for HIV screening at ANC until their onward transition into PMTCT services [26]. This can be explained by the higher cost of equipment and their maintenance, in addition to relatively expensive experienced experts who had to manage this equipment. The higher costs of infrastructure and other inputs into HIV screening at ANC in some of the selected health facilities meant that more patient visits resulted in no reduction of cost per PPY and this yielded no economies of scale. In fact, previous studies have also used the same argument to explain cost differences [27].

In this study the HIV/Syphilis combo was the most costly test kit at USD 3.14 per test kit followed by HCV test kit and HBV at USD 2.47 and USD 0.28 respectively. The Test kit costs dominated overall service costs, representing 90-98% of total per-visit costs depending on test type and health facility. There was no noticeable difference in the cost of the test kits between the Intervention and Control groups respectively. This is because the procurement was done once and the kits were distributed to the study sites at the same time prior to the start of data collection. These findings are comparable to those in a study conducted in Tanzania where procurement of laboratory reagents, RDTs, and other medical consumables was also centrally done [28]. It was established that laboratory supplies and consumables were noted to have claimed a very large share of recurrent/variable cost. It constituted approximately 85% of the recurrent costs incurred in the study [28]. Findings from this study and the Tanzanian study imply that in order to scale up blood born infectious diseases screening using the Integrated Care Model, planning for, and procuring adequate laboratory supplies including RDTs and consumables is important. This is particularly so where a scale up of uptake of screening services for HBV, HCV and HIV targeting pregnant women in first trimester is recommended.

In this study it was established that the CCR for facility space averaged USD 0.0256 per minute (±0.0066). Thus, exhibiting moderate variation across facilities, ranging from USD 0.01593 to USD 0.03474 per minute. Space costs varied considerably across facilities. Lobule HCIII in the Intervention Group demonstrated the highest space CCR at USD 0.03474 per minute, more than twice that of Kijomoro HCIII in the Control Group (USD 0.01593/min), the lowest-cost facility. Since the sample had only HC IIIs, it was noted that there is moderate variation in space CCRs across facilities. Larger facilities, for instance; HCIII with larger ANC rooms, had lower CCRs, indicating economies of scale, as compared to those with limited space or high maintenance costs which had higher CCRs. The existing variations in the CCRs for space within the Intervention Group and the Control Group could be attributed to differences in space utilisation rates. For instance, how many hours per day the rooms are actively used, building/room size, and building maintenance costs. Larger ANC rooms demonstrated economies of scale, resulting in lower space CCRs. Similarly, in a study conducted in Uganda on Costs and resource distribution of direct services for HIV found a space CCR of less than 1% [10]. This cost distribution aligns with ABC/M analyses study conducted in Tanzania, as well as prior research by others in Uganda conducted [21-29]. While local context costing evidence is relevant for healthcare planning, budgeting and cost-effectiveness analysis, and especially on integration of infectious diseases (HBV, HCV and HIV) it continues to be scarce in Uganda. This study is the first to evaluate integration of HBV, HCV and HIV screening, and also to estimate cost of screening at one point of care (ANC) using the TDABC Model in Uganda, and probably in East, Central horn of Africa.

## Limitations of the study

Findings from this study are limited first by the data perspective used in this study; The providers’ perspective may not provide a holistic view of costs as would, the participant’s perspective [9]. The assumptions used in the costing are only applicable at the time of the study as costs are constantly changing. Therefore, these findings should be interpreted based on the assumptions made at the time of the study.

To the best of our knowledge (strengths); Our study is the first to establish the costs of integrated HBV, HCV and HIV screening pathways using ICM in Eastern, Central and Horn of Africa using primary data from a facility-based interventions compared to standard care package. The available economic evaluations of HBV, and HIV interventions in low-income and middle-income countries did not look at integrated screening, rather treatment, HIV integration and other prevention of HBV infection through vaccination [22–24]. This study provides credible evidence and framework for further studies to be conducted. The costing methodology used here can be replicated in a similar setting with a high HBV and HCV burden.

## Conclusion

Overall, the integration of HBV, HCV, and HIV screening using the Integrated Care Model that this study piloted resulted in:

Cost efficiencies where the same staff and space were used for multiple simultaneous tests. Reduced marginal costs for HIV tests due to larger procurement volumes. Higher marginal cost additions for HBV and HCV due to pricier RDTs and reagents

Variations in cost of screening and care at ANC highlight that there may be opportunities for quality improvement, reduced bottlenecks, and reduced waiting times through standardizing care using the IMC for instance, and determining the ideal flow of patients at a health facility.

Health facility-based testing of infectious diseases targeting reduction of vertical transmission among women at one-point-of care such as ANC, and those with similar route of transmission such as HBV, HCV and HIV remains an essential service to meet universal access goals and enhance better health outcomes for the mothers and newborns.

**Annex 1: Sample Process Maps developed in this study *Figure 1 & 2***

**Figure 1:**
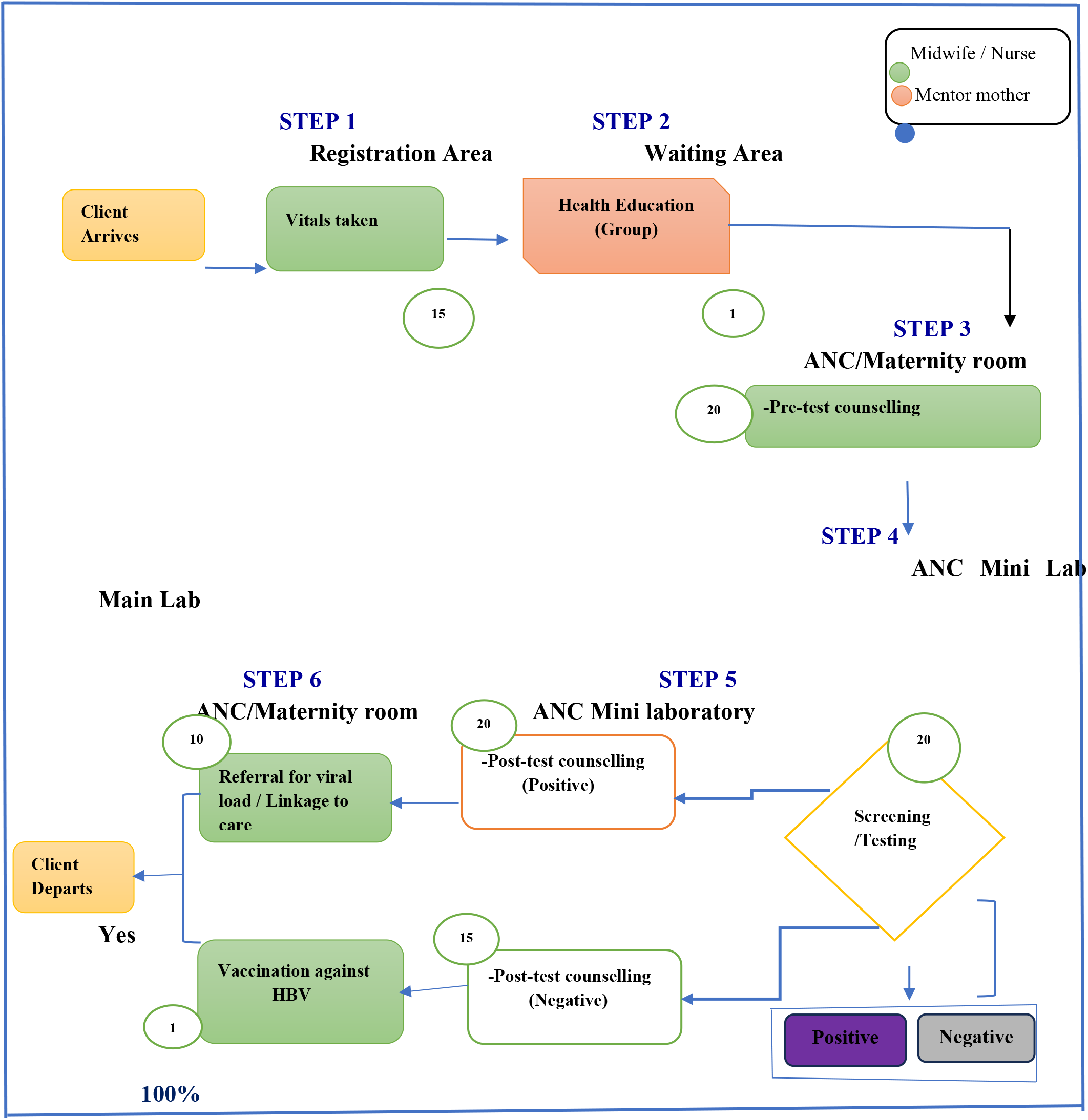
Sample Process Maps in the Intervention Group Ayipe HC III, Koboko District. **Source:** Process map constructed by the PI (2025)

**Figure 2:**
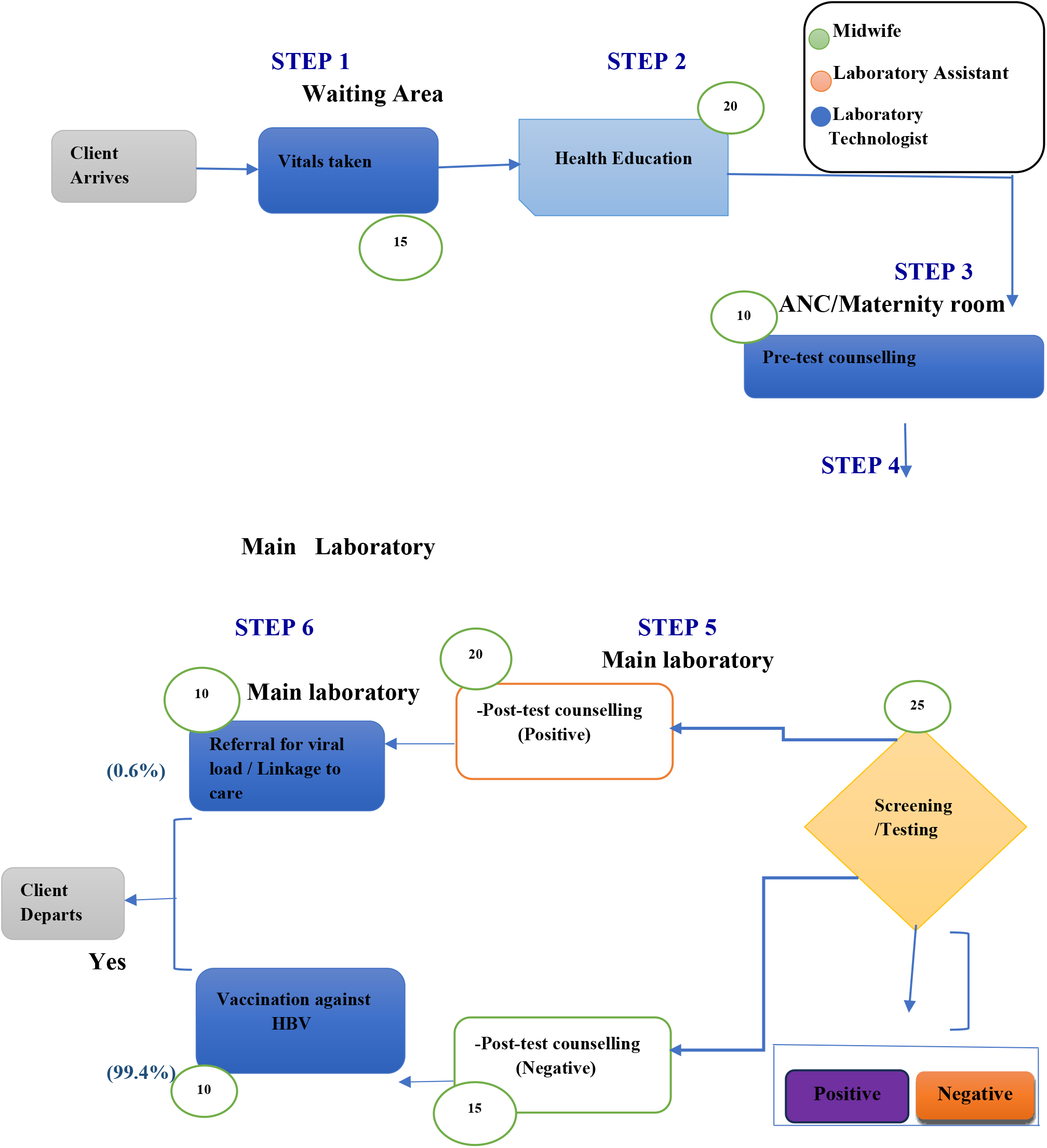
Sample Process Map in the Control Group Eliofe HC III, Maracha District. **Source:** Process map constructed by the PI (2025)

## Data Availability

No legal or ethical restrictions

## Acknowledgements

The authors would like to acknowledge Lugei Foundation, SD-Biosensor for the support. Clarke International University, and Nairobi, Kenya, to which the authors are affiliated, and their support throughout this research project.

## Author contributions

Conceptualized the study, developed the research protocol, collected data, and wrote the first draft of the manuscript: Jon Bosco Alege

Provided technical support from conceptualization to the finalization of the research project: Alloys S.S Orago Provided all the guidance regarding compliance, quality check, and overall review of the research report and manuscript; John Paul Oyore.

Field supervisor, and also provided the oversight and conducted technical review of the manuscript; Philippa Musoke

Jointly provided guidance in the methodology, cleaned and analyzed the data: Anthony Ssebagereka and Richard Ssempala.

## Data Availability Statement: Data availability statement

All relevant data underlying the findings of this study are included in the manuscript and its Supporting information files (Datasets1 & 2).

## References

1. UNAIDS. Global HIV & AIDS statistics– 2020 fact sheet. Available from: https://www.unaids.org/en/resources/fact-sheet. Accessed August 6, 2025.

2. Jefferies M, Rauff B, Rashid H, Lam T, Rafiq S. Update on global epidemiology of viral hepatitis and preventive strategies. World J Clin Cases. 2018;6 (13):589. doi:10.12998/wjcc.v6.i13.5894.

3. World Health Organization (WHO). Global hepatitis report 2017. World Health Organization; 2017.

4. Liz Highleyman, 2017. Hepatitis B and HIV Available at: Hepatitis B and HIV | aidsmap

5. Church K, Mayhew SH. Integration of STI and HIV prevention, care, and treatment into family planning services: a review of the literature. Stud Fam Plann. 2009 Sep;40(3):171–86. doi: 10.1111/j.1728-4465.2009.00201.x. PMID: 19852408.

6. Zhang, Lei, et al. “Integrated approach for triple elimination of mother-to-child transmission of HIV, hepatitis B and syphilis is highly effective and cost-effective: an economic evaluation.” International journal of epidemiology 48.4 (2019): 1327–1339.

7. Job F. H. Eijsink et al., (2020) Cost-effectiveness of hepatitis C virus screening, and subsequent monitoring or treatment among pregnant women in the Netherlands. The European Journal of Health Economics (2021) 22:75–88 10.1007/s10198-020-01236-2

8. Obure, Carol Dayo, et al. “The costs of delivering integrated HIV and sexual reproductive health services in limited resource settings.” PloS one 10.5 (2015): e0124476.

9. Ejalu DL, Mutyoba JN, Wandera C, Seremba E, Kambugu A, Muganzi A, Beyagira R, Amandua J, Mugagga K, Easterbrook P, Ocama P. Integrating hepatitis B care and treatment with existing HIV services is possible: cost of integrated HIV and hepatitis B treatment in a low-resource setting: a cross-sectional hospital-based cost-minimisation assessment. BMJ Open. 2022 Jul 1;12(7):e058722. doi: 10.1136/bmjopen-2021-058722. PMID: 35777868; PMCID: PMC9252200.

10. Ekirapa E, Jordan M, Nong T, et al. (2024). Costs and resource distribution of direct services for HIV in Uganda. BMJ Open 2024;14: e082062. doi:10.1136/bmjopen-2023-082062

11. Robert S. Kaplan Steven R. Andrrson (2004). Time-Driven Activity-Based Costing (TDABC) Harvard Reviews

12. Robert S. Kaplan Steven R. Andrrson (2007). Time-Driven Activity-Based Costing (TDABC)-A Simpler and more Powerful Path to Higher Profits. Havard Business School Publishing Corporation

13. Hoozée S. & Bruggeman, W. (2010). “Identifying Operational Improvements During the Design Process of a Time-driven ABC System: The Role of Collective Worker Participation and Leadership Style”. Management Accounting Research, 21(3), 185–198.

14. Lumbwe Chola, Ryan McBain and Y-Ling Chi (2022), Costing Healthcare Services Using Time-Driven Activity-Based Costing: A Simple Step-By-Step Guide for Data Collection and Analysis. CGD Policy Paper 271. Washington, DC: Center for Global Development. [Accessed on 17th August, 2025]. Available at: https://www.cgdev.org/publication/costing-healthcare-services-using-time-driven-activity-based-costing-simple-step-ste

15. Thin K, Prum V, Johns B (2019) The cost of HIV services at health facilities in Cambodia. PLoS ONE 14(5): e0216774. https://doi.org/0.1371/journal.pone.021677

16. MoH Mozambique (2022). Applying Activity-Based Costing and Management to HIV Services in Mozambique, MoH, Mozambique.

17. Ekirapa E, Jordan M, Nong T, et al. (2024). Costs and resource distribution of direct services for HIV in Uganda. BMJ Open 2024;14: e082062. doi:10.1136/bmjopen-2023-082062

18. GHCC (Global Health Cost Consortium). 2017. “Reference Case for Estimating the Costs of Global Health Services and Interventions.” Vassall A, Sweeney S, Kahn JG, Gomez GB, Bollinger LA, Marseille E, Herzel B, DeCormier Plosky W, Cunnama L, Sinanovic E, Bautista-Arredondo S, GHCC Technical Advisory Group, GHCC Stakeholder Group, Harris K, Levin C. https://ghcosting.org/pages/standards/reference_case.

19. Vyas Seema et al., (2021). Cost variations in prevention of mother-to-child HIV transmission services integrated within maternal and child health services in rural Tanzania. GLOBAL PUBLIC HEALTH2021, VOL. 16, NO. 2, 305–318 10.1080/17441692.2020.1798486

20. Bautista-Arredondo, S., Sosa-Rubí, S. G., Opuni, M., Contreras-Loya, D., Kwan, A., Chaumont, C., Chompolola, A.,Condo, J., Galarraga, O., Martinson, N., Masiye, F., Nsanzimana, S., Ochoa-Moreno, I., Wamai, R., & Wang’ombe, J. (2016). Costs along the service cascades for HIV testing and counselling and prevention of mother-to-child trans-mission. AIDS, 30(16), 2495–2504. 10.1097/QAD.0000000000001208

21. Vyas Seema et al., (2021). Cost variations in prevention of mother-to-child HIV transmission services integrated within maternal and child health services in rural Tanzania. GLOBAL PUBLIC HEALTH2021, VOL. 16, NO. 2, 305–318 10.1080/17441692.2020.1798486

22. Gosset A, Drabo S, Carrieri P, et al., (2024) Costs of integrating hepatitis B screening and antiviral prophylaxis into routine antenatal care in Burkina Faso: Treat all versus targeted strategies. Int J Gynecol Obstet. 2024; 166:44–61. doi:10.1002/ijgo.1551

23. Mwenge L, Sande L, Mangenah C, Ahmed N, Kanema S, d’Elbee M, et al. (2017) Costs of facility-based HIV testing in Malawi, Zambia and Zimbabwe. PLoS ONE 12(10): e0185740. 10.1371/journal.pone.0185740

24. Yigezu et al, (2020). Cost-effectiveness of facility-based, stand-alone and mobile-based voluntary counselling and testing for HIV in Addis Ababa, Ethiopia Cost Eff Resour Alloc (2020) 18:34 10.1186/s12962-020-00231-x

25. Sabin L, Haghparast-Bidgoli H, Thapaliya B, Chand O, Bhattarai S, Arjyal A, et al. (2024) Factors influencing the implementation of integrated screening for HIV, syphilis, and hepatitis B for pregnant women in Nepal: A qualitative study. PLOS Glob Public Health 4(10): e0003006. 10.1371/journal.pgph.0003006

26. Zegeye et al, (2019). Assessing the cost of providing a prevention of mother-to-child transmission of HIV/AIDS service in Ethiopia: urban-rural health facilities setting BMC Health Services Research (2019) 19:148 10.1186/s12913-019-3978-4

27. Ahmed, N., Ong, J.J., McGee, K. et al. Costs of HIV testing services in sub-Saharan Africa: a systematic literature review. BMC Infect Dis 22 (Suppl 1), 980 (2022). 10.1186/s12879-024-09770-7

28. Kahwa et al., (2008). Exploring the Costs of Laboratory Testing for HIV and Major Sources of Funding for Tanzania. Research Journal of Medical Sciences 2 (1): 33–37, 2008.

29. Aggrey D. Mukose et al., (2020). Costs and Cost Drivers of Providing Option B+ Services to Mother-Baby Pairs for PMTCT of HIV in Health Centre IV Facilities in Jinja District, Uganda. Hindawi BioMed Research International Volume 2020, Article ID 2875864, 9 pages 10.1155/2020/2875864

30. Husereau D, Drummond M, Augustovski F, de Bekker-Grob E, Briggs AH, Carswell C, Caulley L, Chaiyakunapruk N, Greenberg D, Loder E, Mauskopf J, Mullins CD, Petrou S, Pwu RF, Staniszewska S; CHEERS 2022 ISPOR Good Research Practices Task Force. Consolidated Health Economic Evaluation Reporting Standards 2022 (CHEERS 2022) Statement: Updated Reporting Guidance for Health Economic Evaluations. Value Health. 2022 Jan;25(1):3–9. doi: 10.1016/j.jval.2021.11.1351. PMID: 35031096.

